# In Holistic Admissions, a Combination of Non-Academic Explanatory Variables Has Significant Predictive Value for Applicant Ranking

**DOI:** 10.1101/2024.11.01.24316296

**Authors:** Andrew D. Bergemann, Stephen R. Smith, Joel A. Daboub

**Author notes:** Dr. Bergemann was an Associate Professor in the Department of Medical Education at the Dell Medical School, at the University of Texas at Austin, Austin, Texas, USA during the concept development, data collection, and analysis phases of the study. Manuscript preparation was performed after Dr. Bergemann moved to Baylor College of Medicine.

## Abstract

A medical school with a focus on community engagement has innovated its admissions process to include three different interview formats and one novel task format. Each component is designed to assess specific attributes of applicants, including teamwork skills, cultural competence, and service orientation. Correlations between these components are low, consistent with the original purpose that each should assess different attributes. To understand the use of the data by the members of the committee that ranks applicants, the authors created a model of seven explanatory variables, comprised of the three interview ratings and one task rating, a review of the written applications, and two measures of past academic performance. With regression analysis, the model significantly predicted applicant rankings, with most of the predictive capacity retained after omission of academic metrics. The results display that the school has developed innovations that allow for a reduced dependence upon academic history, and instead uses a truly holistic approach that is tailored to its mission. Most importantly, the work establishes that the admissions committee uses all the diverse forms of data provided to make decisions, which until this point has been an open question in holistic admissions.

## Introduction

Holistic review allows for the evaluation of potential candidates based on the totality of their activities, characteristics, and lived experiences as demonstrated up to the point of their application for admission. This type of review provides evidence of a candidate’s commitment to the practice of medicine and to caring for those who are ill, as well as how they themselves overcome adversity or barriers in their own lives and how they learn from such experiences.

The benefits of healthcare professionals reflecting the demographic and cultural composition of the communities that they serve are well established [1–3]. In the Southern United States, a clear example of the need for better demographic representation is the geographic under-representation of rural communities amongst clinicians, contributing to these communities being under-served for healthcare [4–8].

Greater inclusion of many under-represented groups begins with their acceptance into health-related professional schools. Society has decided that this cannot happen through race-based admissions processes, presenting a challenge as communities of color are often amongst the most under-represented in the healthcare professions and experiencing the most barriers to healthcare access. Ample evidence demonstrates that the use of holistic admissions allows for the recruitment of candidates with desirable qualities, alters the demographic composition of matriculating classes (frequently increasing diversity without race-based affirmative action), and can also at least sometimes exceed past academic performance as a predictor of future academic or career success [9–12]. Holistic admissions, therefore, has the capacity to enable the entry into professional schools of more applicants from under-represented groups, without that demographic change being the goal of the process, but rather by recognizing individual candidates for their non-academic strengths.

A school’s admissions processes should be tailored to fit its mission and teaching philosophies [13]. We report here on an admissions process at a Southern United States medical school that emphasizes innovation, leadership and community service and that teaches using teamwork environments [14–16]. To facilitate the search to identify applicants who prioritize the school’s mission values, and who will excel in the school’s learning environment, the school initiated a number of innovative approaches to rate applicants. Most notably, the school initiated a task-based assessment, which the applicants accomplish in teams. During the performance of the task, applicants are assessed for their teamwork skills [14].

The school also uses written application reviews, asynchronous video interviews, multiple mini-interviews (MMIs), and classical interviews, all of which requires a significant commitment of staff and faculty time [14]. The video interviews include questions tailored to address an applicant’s past history of innovation, community impact, and experiences relevant to teamwork and leadership. The MMIs focus on the capacity of each applicant to address complex situations with empathy for stakeholders with conflicting interests. Interviewers in the classical interview format are trained to focus on each applicant’s communication skills and their experiences, particularly those reflecting cultural competence. Due to the extensive nature of the process, considerable resources are required to complete all the components. Some importance therefore must be placed on whether the final rankers of the applicants, the members of the Admission Selection Committee (ASC), use the data from all the rating mechanisms.

While many studies establish that holistic admissions has positive outcomes, relatively few professional schools have sought to identify the relative impact of the components of their admissions processes, with two notable exceptions [17,18]. For those two studies, the authors use broader categories of explanatory variables. Hence, our research begins to fill a gap in the existing literature that is central to holistic admissions, at a time when it is spreading widely through higher education.

Specifically, in this research, we establish that all of the school’s ratings contribute significantly to a predictive, multiple linear regression (MLR) model of applicant ranking, consistent with a holistic process, in which final rankers use all the applicant data provided to them.

## Materials and Methods

Data was collected for the 2018 (recruiting the 2019 matriculating class) and the 2019 (recruiting the 2020 matriculating class) admissions seasons. These two years were chosen as the admissions process was consistent across these two years. Data collection began 5/1/2018 and continued through to 2/1/2020. This data collection was the normal collection that occurred for the purposes of admissions, including applicant selection and program assessment and development. Once we had decided to publicly share the conclusions, data was accessed for the purpose of analysis for the study in this report on 6/3/2022.

During the review of applicant’s applications, evaluators were trained to review each written application for evidence of attributes that align with mission. These attributes included “Integrity and Ethics”, “Social and Interpersonal Skills”, “Resilience and Adaptability”, and “Cultural Competence”. The raters were further asked to evaluate candidate’s applications for experiences that were aligned to the mission, including evidence of “Teamwork Experiences”, “Service Orientation”, and the “Development and Implementation of Innovative Solutions”. Evidence of these attributes and experiences was discernible in each applicant’s essays, awards, and lists of experiences. It is important to note that this evaluation of written applications was completed in the absence of academic credentials and letters of recommendation.

Candidates under consideration for interview, based on reviews of their written applications, were invited to an asynchronous video interview. The asynchronous video interview has many parallels to MMIs, and seeks to specifically assess applicants for past activities demonstrating leadership, teamwork, community actions, and innovation. Data from the video interview, combined with the ratings of the written application, determined whether a candidate would be invited to visit the school for an interview day. Interview days included one classical interview, similar to the interviews that have been the mainstay of medical school admissions for decades, and five MMIs [19,20]. Additionally, interview days included a group task, in which applicants were assessed for teamwork skills [14]. Numeric scores were assigned to various aspects of performance for each of the interviews and tasks by trained raters. Raters of the group task and the classical-format interviewers were faculty, and mostly not members of the ASC. MMIs were conducted by medical students, faculty, or staff members, again most of whom were not members of the ASC. Data was collected for all 752 applicants who were interviewed over the two years. The data for 729 of the applicants was used for this research as 23 applicants experienced parallel processes, in some cases because they chose to not do the asynchronous video interview (and instead did a written secondary essay). All research reported here was approved by the UT Austin Institutional Review Board (IRB) (STUDY00002235). Informed consent was not deemed necessary for this retrospective study of data collected as part of the normal educational process. Due to the sensitive nature of admissions data, the underlying data for this article cannot be made publicly available. Data was collected and de-identified by one author, and then analyzed by a second author who was blinded to participant identities.

Rankings of candidates were performed by members of the ASC. The committee of twelve to twenty members evaluated the files of every interviewed candidate. The total applicant evaluation process resulted in over sixty points of evaluation for each candidate, each of which was associated back to an aspect of the mission or curriculum. To manage consistency of the selection process, the data for each interviewed candidate was presented to the committee in an aggregated heatmap that facilitated an overall comparison of candidate evaluations (Fig 1). Each of the structured evaluations were grouped in categories: “Desirable Attributes”, “Experiences”, “Mission Alignment”, and “Communications”. This evaluation was further enhanced by the color gradient that aligned with performance on the rubric and that provided ASC members additional clarity and depth to the metric’s presentation. By associating specific colors with different levels of performance, the information was more visually intuitive and accessible. Adding a visual representation enhanced understanding and allowed committee members to quickly grasp each applicant’s performance across various criteria.

**Figure 1.**
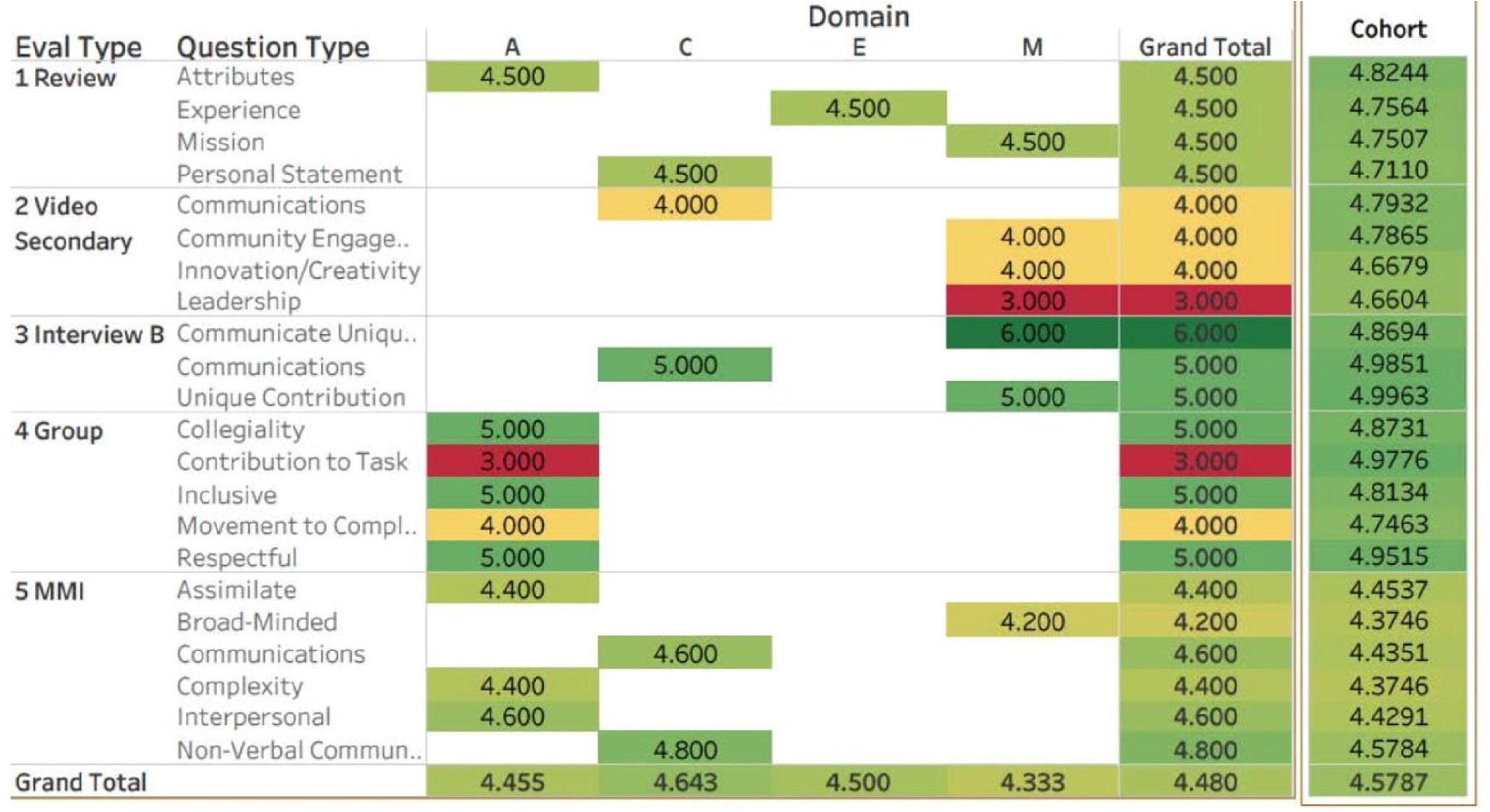
Schematic form for presentation of an applicant to ASC members. This figure displays the typical format for presenting an applicant’s information as a heat map to ASC members. The heat map is coded red to yellow to green equating to poor to reasonable to strong performance in a rating. This format, or only very minor variations on this format, has been used for all interviewed applicants since 2016. Ratings are grouped as being relevant to attributes (A), communication skills (C), experiences (E), and mission fit (M). “1 Review” refers to the ratings of the written application. “2 Video secondary” refers to the asynchronous video interviews. “3 Interview B” refers to the classical one-on-one interviews. “4 Group” refers to the ratings of the group task. “5 MMI” refers to the average ratings across five multiple, mini-interviews. In addition to the heat map, for each candidate the ASC members are also presented with the complete original written application (including letters of recommendation), and any narrative comments provided by application, interview, or task raters.

For each applicant, three ASC members were assigned the role of being primary reviewers, and reviewed in detail all aspects of the candidate’s application and interview performances, and assigned an initial rating of reject, accept tier three, accept tier two, or accept tier one. For applicants with a unanimous rating, independently determined by all three primary reviewers, the rating was automatically confirmed by vote without further discussion. For the majority of applicants, with primary reviewer scores that were not unanimous, the three primary reviewers presented each candidate to the committee for discussion, with at least eleven committee members present. After the discussion, all present members of the committee voted on the candidate. Committee members also voted reject, accept tier three, accept tier two or accept tier one. These votes were then translated to scores: reject: 0; accept tier three: 1; accept tier two: 2; accept tier one: 3. For each candidate the scores from all ASC members were averaged to derive the admissions ranking score used in the following analyses. The choice of this average as the outcome variable of our modelling reflected the fact that this metric also determined a candidate’s selection for an invite to matriculate at the school.

In order to determine the contribution of different components of the process to final applicant ratings, correlational analyses and multiple linear regressions (MLRs) were conducted in R, using the functions *ggpairs* (in *ggplot2* package), *lm* and *summary.lm* (both in *stats* package), broadly as has been described elsewhere [21]. Explanatory variables for past academic history were the GPA and MCAT of each applicant, expressed as standard deviations from the mean in the interviewed cohort. Explanatory variables for non-academic components of the data were the average ratings for each mechanism of the process (written application ratings, video interview ratings, classical interview ratings, MMI ratings, and group task ratings). Appropriateness of the use of multiple linear regression (MLR) was assessed using the R functions *plot* and *augment* (in *broom* package), and *coeftest* and *bptest* (both in *lmtest* package), with results reproduced in *ggplot* (in *ggplot2* package) for publication. Hierarchical MLR was performed using *anova* in R, as described elsewhere [22].

## Results

As a first step to identifying explanatory variables for use in a regression analysis, we tested a series of seven potential predictors of admissions outcomes through determining the correlation of each predictor with the final admissions ranking score (Fig 2). In correlational analyses, each predictor (that is independent or explanatory variable) yielded a positive correlation with the dependent variable between 0.17 and 0.502 (Fig 2).

**Figure 2.**
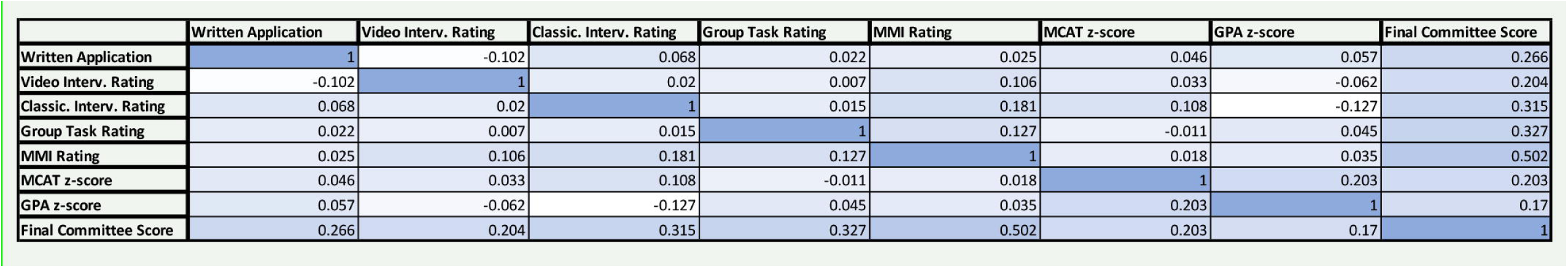
Correlation (R) of explanatory variables with each other and with the dependent variable. Table shows correlations of the seven explanatory variables with each other and with the dependent variable. The seven explanatory variables are the average rater evaluation of each student’s written application, video interviews, classical interview, group task performance, MMI performance (all scored on a range of 1 to 6), the MCAT score and the GPA (both reported as z-score within the interviewed cohort). The dependent variable is the final committee ranking score, calculated as described in the methods section.

MLR revealed that a model (described here as model one) based on the seven predictors yields an R square of 0.52, with a strong likelihood of significance (p<10^−15^ for the model as a whole) (Table 1). The least supported individual explanatory variable was the applicants grade point average (GPA), which was still highly significant (p<10^−6^) and still displayed a strong effect size (expressed as a standardized regression coefficient). Model refinement based upon an adjusted R square approach supported retention of all explanatory variables (data not shown).

**Table 1.** Multiple linear regression predictive models of admissions outcomes.

|  | Model One |  |  |  |  |  |
| --- | --- | --- | --- | --- | --- | --- |
|  | Estimate | Std. Error | t value | Pr(>\|t\|) | Stdev. of Variable | Effect Size |
| **Intercept** | -4.889 | 0.276 | -17.711 | < 2e-16 |  |  |
| **Written App. Review** | 0.311 | 0.034 | 9.034 | < 2e-16 | 0.569 | 0.235 |
| **Video Interview Rating** | 0.164 | 0.024 | 6.972 | 7e-12 | 0.837 | 0.182 |
| **Classical Interview Rating** | 0.216 | 0.022 | 9.714 | < 2e-16 | 0.903 | 0.259 |
| **Group Task Rating** | 0.231 | 0.023 | 10.144 | < 2e-16 | 0.86 | 0.264 |
| **MMI Rating** | 0.556 | 0.038 | 14.626 | < 2e-16 | 0.52 | 0.388 |
| **MCAT z-score** | 0.137 | 0.02 | 6.864 | 1.1e-11 | 1 | 0.181 |
| **GPA z-score** | 0.104 | 0.02 | 5.196 | 2.65 e-7 | 1 | 0.138 |
| **Model Rsquare** | 0.523 |  |  |  |  |  |
| **Adjusted Rsquare** | 0.518 |  |  |  |  |  |
|  | Model Two |  |  |  |  |  |
|  | Estimate | Std. Error | t value | Pr(>\|t\|) | Stdev. of Variable | Effect Size |
| **Intercept** | -4.935 | 0.238 | -20.779 | < 2e-16 |  |  |
| **Written App. Review** | 0.31 | 0.029 | 10.579 | < 2e-16 | 0.569 | 0.256 |
| **Video Interview Rating** | 0.19 | 0.02 | 9.348 | < 2e-16 | 0.817 | 0.255 |
| **Classical Interview Rating** | 0.234 | 0.02 | 11.919 | < 2e-16 | 0.871 | 0.295 |
| **Group Task Rating** | 0.233 | 0.02 | 11.541 | < 2e-16 | 0.823 | 0.278 |
| **MMI Rating** | 0.521 | 0.032 | 16.08 | < 2e-16 | 0.526 | 0.397 |
| **MCAT z-score** | 0.127 | 0.017 | 7.458 | 3e-13 | 0.99 | 0.182 |
| **GPA z-score** | 0.124 | 0.018 | 7.105 | 3.06-12 | 0.975 | 0.175 |
| **Model Rsquare** | 0.614 |  |  |  |  |  |
| **Adjusted Rsquare** | 0.61 |  |  |  |  |  |

Model one describes the analysis of a model incorporating all seven proposed explanatory variables applied to the full cohort of applicants. Model two, also uses all seven explanatory variables, but applied only to the retained cohort after forty-two outliers have been removed, as described in text. Estimate: the coefficient for each variable; Written App. Review: rating of the written application; Stdev of Variable: standard deviation of the explanatory variable; Effect Size: the number of standard deviations change in the outcome variable (average committee rating) predicted as a response to a one standard deviation change in the explanatory variable.

The potency of the written application, task, and interview ratings upon candidate selection is substantial. The effect of each of these variables upon the committee average rating is described in Table 1 as the effect size, measured as the predicted change of the outcome variable (in standard deviations), resulting from a one standard deviation change in an explanatory variable, all else being held constant. For each of the written application, task, and interview ratings (that is asynchronous video interview, MMI, and classical interview), the effect size is in the range of 0.18 to 0.40. As an example, all else being held constant, a student increasing their group task performance by one standard deviation would be predicted by model one to improve their admissions ratings score by 0.264 standard deviations. Clearly this represents change at the level at which the interview ratings or task performance ratings can substantially change an applicant’s likelihood of receiving an enrollment offer.

It is worth noting that each of the non-academic history metrics are equally or more potent than either of the academic history metrics (that is GPA and MCAT). Of the explanatory variables, the MMI provides the greatest contribution to predictive power, indicating potentially that it also has the most influence on ASC members. It is interesting to consider that ASC members may be making a rational choice, recognizing that the MMI scores represent data from five different encounters and five different raters for each applicant.

A series of tests need to be performed to determine the appropriateness of a dataset, and the inferred model, for analysis by multiple linear regression. First, is to test whether the independent variables display collinearity. While collinearity does not preclude regression analysis, it does present a significant limitation as it increases the error on the measured coefficients [23,24]. All the independent variables of the model display correlations of less than 0.21 (Fig 2), implying relatively low collinearity. Of note, this result also strongly suggests that our different interview formats assessed different qualities or attributes in candidates, vindicating the investment required to conduct them. We also find that the model meets established criteria for linearity, with mean residual values approximating zero across all fitted (predicted) values (Fig 3A). The data is broadly homoscedastic, with the exception of a small number of individuals at high fitted values (Fig 3E). Of greatest concern is that analysis using the Breusch-Pagan test rejects the hypothesis that the heteroscedasticity is insignificant (p = 0.025). The danger of heteroscedasticity is that it can lead to inaccurately low standard errors, and therefore inaccurately low p value estimates of relationships. To address this concern, we performed the widely used approach of developing robust standard errors [25,26], using the functions *coeftest* in R. The robust standard errors generated for all variables increased by less than 10% from the originals, except for the standard error for the group rating which increased by less than 15%.

**Figure 3.**
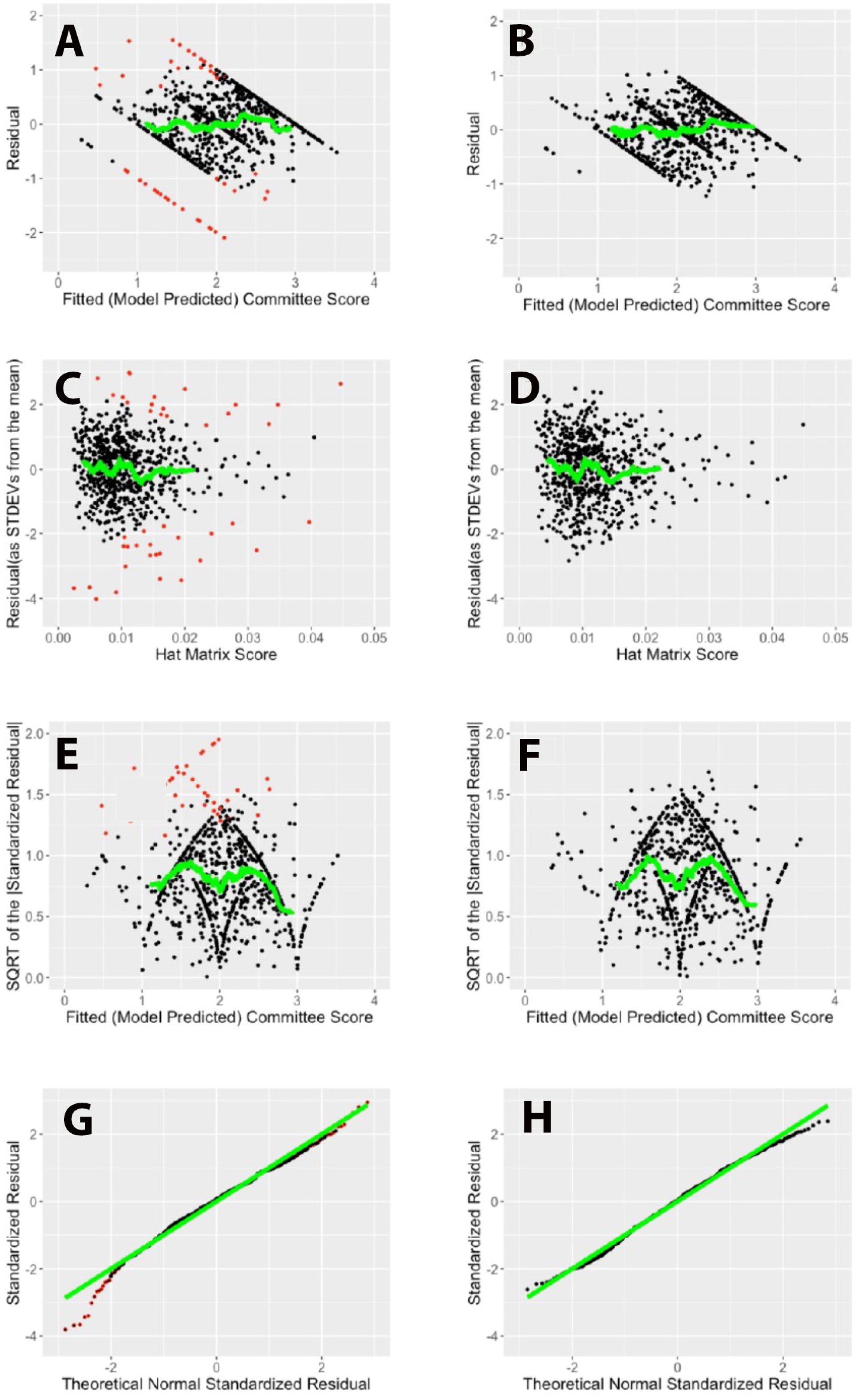
Graphs testing the appropriateness of MLR to test the models. Graphs 3A, 3C, 3E, and 3G represent the model derived from the 2019 and 2020 combined cohort. Graphs 3B, 3D, 3F, and 3H represent the second model, derived from the original cohort after removal of influential and extreme outliers. Graphs 3A and 3B depict the relationship of each applicant’s residual (the difference between their actual score and their fitted score) (y axis), and their fitted score (x axis). Ideally model linearity is represented by a horizontal line at a y value of 0. Both models approximate linearity well. Graphs 3C and 3D depict the relationship of each applicant’s leverage score (as measured by hat matrix) (x-axis) and their standardized residual (y axis). Highly influential scores are those with both high leverage and high residual scores. Graphs 3E and 3F represent the relationship of the square root of the absolute value of each candidate’s residual (y axis) to their fitted score (x axis). Ideally model homoscedasticity is represented by a horizontal line. Both models approximate homoscedasticity except at high fitted scores. Although not obvious from the graphs, the second model has less heteroscedasticity, as measured by the Breusch-Pagan test (for the second model, the test does not reject the null hypothesis, which proposes that there is no heteroscedasticity).

Given the very low p values generated to test the significance of the relationships (Table 1), these small changes in standard error indicate heteroscedasticity does not represent a concern for the validity of the model.

Graphs 3G and 3H depict the relationship of the distribution of each model’s residuals (y axis), and a theoretical normal distribution (x axis). Ideally normality of residuals is represented by a straight line with all points on the line. Both models approximate linearity reasonably well. Graphs 3A, 3B, 3C, 3D, 3E, and 3F display lines (green) indicating a running average centered on a window of 70. Graphs 3G and 3H each show the theoretical line (green) for perfect normality. For graphs 3A, 3C, 3E, and 3G, points representing individuals omitted from the second model are indicated in red.

The residuals (deviation of individual applicant’s outcome score from the model’s fitted score) display reasonable normality except for a small excess of the very negative residuals (Fig 3G). We have chosen to use visualization of normality in a Q-Q plot to assess normality, rather than quantitative tests of normality, as quantitative tests of normality are excessively sensitive to small aberrations with large data sets [27].

Finally, the model was tested for the influence of outliers. The most influential outliers can be calculated using Cooks distance (CD). Forty-one applicants exceeded the recognized standard of having a CD exceeding 4 divided by the population size (n). For this model, 4/n =4/729 or 0.00549. Using CD values exceeding 4/n is not arbitrarily chosen, but rather is a widely used mechanism to identify outliers [21,24,28,29]. To test the effects of outliers on the model, we created a derived population, from which forty-one applicants with the highest CD (that is those with a CD exceeding 0.00549) have been removed, as well as one additional applicant with a very high standardized residual (>3) (the applicants excluded from the new population are those marked in red in the relevant figures) (Figs 3A, 3C, 3E, and 3G). We created a new model using the derived population and the same seven explanatory variables. The new model (model 2) modestly out-performed the original (Table 1, and Fig 3), with an increased R square (R square equals 0.61), a non-significant Breusch-Pagan statistic (p=0.29), and a slightly improved normality of the residuals (Fig 3H).

The best approach to accommodating influential outliers in regression analyses has been a constant debate in statistics, with valid arguments for both their retention and omission from analyses [30,31]. We have taken what we believe to be the most accurate and ethical approach in this research, describing our results before and after omission. However, we recognize that many researchers take the position that outliers should only be removed when there is a compelling justification. In our case, we believe the compelling justification is that the outliers do seem to represent a separate sub-population, as 16 of the 42 outliers who were excluded from the derived population were ranked as “reject” by the committee. Of the 687 applicants in the derived population, only five were ranked as reject. The result suggests that major determinants for decision of “reject” fall outside of the seven explanatory variables used in the models. Indeed, we are aware that “reject” status is frequently prompted by an applicant’s history of legal problems or academic malfeasance. We have not attempted to incorporate these potential explanatory variables as they are both hard to numerically describe and applicable to only a small fraction of applicants. Removal of the influential data points only modestly affected our model metrics (Table 1), indicating the robustness of the models. The relative constancy of the metrics also strongly suggests that the modest heteroscedasticity present in model one has little effect on the model’s predictive value. Importantly, while we consider that the results after removal of outliers are interesting, the correlations are clearly present and validated when the outliers are included. Hence, our conclusions are not dependent on the removal of the outliers.

Not all of the seven explanatory variables used in the models are normally distributed. In particular, the z scores of GPAs are highly left-skewed. However, most statisticians agree that skewing of explanatory variables does not represent a problem for linear regression [32,33].

It is interesting to consider the sources of the unaccounted variance. As none of the model’s explanatory variables include an assessment of applicants’ letters of recommendation (LORs), the LORs represent a source of unaccounted variance. The members of the ASC have access to the LORs, unlike any of the raters of the written applications, interviews, or tasks. It is also possible that the model deals poorly with truly exceptional experiences such as receiving a major international award for humanitarianism, or having a history of academic probation, or legal problems. These highly influential metrics likely skew the committee’s decision away from the model’s fitted value for an individual.

It is notable that a more limited model that includes only our task, interview (all three formats), and written application assessments as explanatory variables still retained significant predictive value, with an R square of 0.46. As task, interview, and written application raters never saw the student’s MCAT, GPA, or letters of recommendation, the more-restrictive model establishes that ASC members are influenced by factors external to academic history. In other words, this result is a validation that the process is holistic. In fact, using a hierarchical approach to the regression analysis, beginning with a model including just academic metrics, and then adding in non-academic metrics, the substantial effect of the non-academic metrics is made clear. A model with just MCAT and GPA as variables yields an R square of 0.05, compared to the R square of 0.52 once the non-academic metrics are added. The change in R square from introducing the non-academic ratings is 0.47 and is statistically significant by F-test (F(5, 721)= 140.3; p<10^−15^).

The null hypothesis for this study is that admissions committee ratings, and therefore, offers for admission, are solely correlated with past academic performance, and do not correlate with application, interview, or task ratings. Clearly, the results reject the null hypothesis.

A limitation of our study is that it is correlational and does not directly address causation. Although we believe it is highly unlikely, we acknowledge the possibility that the predictive value of ratings such as the MMI rating and the group task rating may not be due to ASC members using these ratings, but rather due to confounding variables that correlate with these ratings. We acknowledge that the results are very consistent with ASC members using all the data provided to them, rather than a proof of a causative relationship. While establishing a causative relationship would be preferable, study designs that would allow for a determination of causation would be both ethically and practically very challenging.

One might propose that one possible confounding variable could be the letters of recommendation (LORs), if the quality of LORs correlated with the explanatory variables of the models. However, this proposal has limited applicability, as the model variables do not strongly correlate with each other, and hence the LORs cannot confound for multiple components. Indeed, the lack of collinearity among the explanatory variables strongly contributes to our assertion that it is unlikely that any single confounding variable could account for much of the observed predictive value of the models.

A further consideration regarding this work is that each of the authors have at times served as interview raters and as ASC members. However, this research project was retrospective, and the approach was devised after the data construction and collection period. Hence, the authors were effectively blinded to any influence of the research goals.

## Discussion

The ability for the committee to reflect on the candidate’s past behavior and experiences gave them a window into how the candidate may respond to future adversity, care for others no matter the patient’s station in society and innovate and lead in the future as they navigate their training. Similar to an employer inferring the potential of a future employee based on their previous work experience, holistic admissions allows the admission committee to consider not only the candidate’s academic competencies, but also their passions and interests as evidenced by past activities.

To that end, a structured applicant evaluation process that is aligned with the institutional mission can in fact identify those candidates who have a higher likelihood of sharing values and attributes with that mission. At its inception, the school sought to identify students who aligned with the institution’s mission of improving the quality of, and access to, care for the community it served. Through each step of the evaluation process candidates were evaluated not only on their academic metrics, but their attributes, characteristics, and experiences as demonstrated by their past behaviors and the interview day experience.

Holistic admissions seeks to assess and rank applicants for both their academic record (and inferentially, their academic capabilities) and for attributes broadly termed non-cognitive skills. Non-cognitive skills can broadly be described by the social science constructs of self-directed learning (SDL) [34,35], grit [36], and emotional intelligence (EI) [37], combined with both an applicant’s capacity to innovate and their motivation towards community engagement. Grit has been described as a combination of perseverance and a passion for long term goals [36]. A useful description of EI characterizes it as a combination of self-awareness, self-regulation, motivation, empathy, and social skills [37]. Within the social skills category of EI are skills highly valued by the school in the study, including communication, leadership, and teamwork skills. At least in some settings, these social constructs are predictors of academic or vocational success [38].

As we have indicated, this diverse set of attributes that include SDL, grit, EI, community engagement, and innovation are sometimes called “non-cognitive” or “soft skills” [38]. While we recognize that “non-cognitive skills” is the current widely-used term for all metrics other than academic history, we have strong reservations about the implication that these attributes are not intellectual. The fact that they can be both taught and learned, and that they can impact academic success [38,39], strongly implies that they are cognitive. Nonetheless, in recognition of current terminology, we are using non-cognitive in this discussion to describe these attributes that the school values.

While the medical education community has broadly adopted holistic admissions in order to better identify candidates with these desired non-cognitive attributes, testing of the efficacy of holistic admissions has been recognized as critical [13]. This need stems from both the large commitment of resources required for holistic admissions and from the current legal questions surrounding admissions processes [13]. In particular, optimization of the metrics assessed in holistic admissions processes is considered a research priority [40]. The extent of such research is currently limited, with notable examples at the University of Michigan Medical School [17,18]. While these examples used approaches with some similarities to ours, neither study looked at the relative influence or predictive value of different forms of interviews. Our work presents strong correlations that necessarily constitute a foundational cornerstone of holistic admissions research – namely that decision maker’s choices do correlate with all the data presented to them, and that they do not revert to an over-dependence on the applicants’ academic histories.

However, our research raises serious questions. In particular, what should be the optimal mix of influence of academic history and non-cognitive assessments on rankers of applicants for admissions? A school might be content with the observations reported here of this program, but a more research-derived answer should surely be better. The research to answer such a question represents a novel major endeavor beyond the scope of this study. The answer lies ultimately in the performance of a school’s graduates as doctors, and correlating those performance metrics against the admissions metrics supporting their original admission. However, the complexity of such research is revealed simply through asking the question, what is the appropriate measure of a clinician’s performance? The answer to this question clearly changes depending upon the priorities of the individual (or institution) asking the question. For a medical school that prioritizes community engagement, presumably this requires metrics of the on-going engagement of its graduates.

Due to the innovative nature of the described program, some may question the generalizability of the research in this report to other medical schools. However, the central idea of using MLR to test the relative influences of academic and non-academic factors on admissions decisions can be applied to any medical school. The explanatory variables will change from school to school, but the core idea and its application remains relevant. Perhaps more substantively, the processes reported here for the conduct of an innovative admissions process will be valuable for any school that is considering initiating a mission-focused holistic admissions program. Although the details which change dependent of a school’s mission and values, our work provides a general guide for establishing new programs, and for evaluating aspects of their success.

Although we have presented the lack of collinearity between explanatory variables as principally a necessary factor for optimization of MLR, the lack of collinearity is in-itself an interesting result. The lack of collinearity indicates that each of the task and interview ratings test different attributes in the candidates, supporting the retention of each of these different components. Clearly, this is a strong validation of the school’s original goals in designing the process, as each component was designed to test specific attributes. In the case of the video interviews, the goal was to assess applicants for community engagement, leadership, and innovation. The group task was designed to assess their teamwork skills. The classical interview was specifically crafted to address communication and experiences relevant to cultural competence. As is often the case, the MMIs assessed applicants for comprehension of complex problems, broad-mindedness and inclusivity. These are now concepts central to the mission of many medical schools. The lack of collinearity therefore also speaks to the generalizability of this work, as it represents a significant reason for other schools to invest in this multi-prong approach. In fact, a previous study at a different institution showed low to medium correlations between traditional interview assessments and MMI scores, and inferred that that different interview techniques may more fully characterize the attributes of a candidate [41].

The importance of reducing the influence of standardized tests upon admissions decisions has become recognized as an essential, but not sufficient, step for promoting entry of under-represented groups to a range of disciplines and professions [12,42–44]. Currently, preliminary evidence supports the contention that holistic admissions can increase the matriculation rates for under-represented minorities (URMs) into medical schools [10,45], indicating theses applicants have merits under-appreciated by traditional admissions processes. This presumably allows for recruitment of URMs through their desirable attributes and experiences, thereby meeting current legal standards pertaining to admissions. However, the efficacy of holistic admissions clearly depends on the final rankers use of data outside of the MCAT. In this report, strong evidence is provided that this is true for the particular admissions program in the study. It is of value to note that MCAT scores have previously been demonstrated to be poor predictors of post-graduation clinical performance [46].

As mentioned earlier, survey evidence indicates that the school’s matriculants are oriented towards the school’s mission, including an orientation towards leadership, innovation, and being the drivers of social change [14]. There is also ample anecdotal evidence that the school’s students and graduates are oriented toward community services [47–49]. It is challenging to parse out whether these results represent a product of the selection process or the attraction of mission-oriented students toward a school that emphasizes community engagement. Future directions of research potentially investigate qualitatively the motivations and career paths of graduates as they move through their careers.

Legal decisions are pushing admissions toward merit-based assessments. This study describes a process that allows non-cognitive values to be meaningfully assessed and to carry a weight in the holistic assessment of applicants. Hence an evaluation based on “merit” alone can be influential in admissions decisions even when “merit” is assessed in “non-cognitive” but important performance-based evaluations. While merit might be misconstrued as solely academic achievement, merit should surely also be measured as the capacity to persevere in the face of adversity, to have expended time and resources in meaningful community experiences, and to have developed the empathy to understand differing perspectives.

In summary, we have created a MLR model of an admissions process that is significantly predictive of final applicant rankings. The model’s results are consistent with the school’s admissions process evaluating candidates through considering a wide array of information, and that the admissions process is therefore truly holistic. The described processes, both the administration and the evaluation of the admission program, can act as a model for other professional schools to create their own mission-aligned admissions programs.

## Data Availability

All data that can be released are contained within the manuscript. Due to the sensitive nature of admissions data, data cannot be provided in any greater granularity than in the current manuscript.

## Acknowledgements

The authors would like to express our appreciation of the medical students, staff, faculty, and community members, who have engaged in reviewing applications and rating applicants. We particularly would like to thank the members of the Admissions Selection Committee for their diligence and commitment to excellence. We would also like to thank our colleagues in the Kern National Network for Caring and Character in Medicine for valuable interactions.

## Notes

### Competing Interest Statement

The authors have declared no competing interest.

### Funding Statement

This study did not receive any funding.

### Author Declarations

The Institutional Review Board (IRB) of the University of Texas at Austin's Office of Research Support and Compliance waived ethical approval of this work (IRB ID: STUDY00002235).

